# Metal concentrations in breastmilk of Fuzhou nursing mothers associates with dietary and lifestyle factors

**DOI:** 10.1101/2021.07.27.21261197

**Authors:** Lin Qi, Vic Shao-Chih Chiang, Qiu-Yan Lu, Jing-yuan Wen, Rongxian Xu

**Author notes:** **Corresponding author:** Professor Rongxian Xu, 64 2108516764, Address: No.1 Xue Yuan Road, Shang Jie Town, Min Hou, Fuzhou, China.

## Abstract

Metal intake greatly influences physiological development during infancy. For infants, breastmilk is the major route of metal exposure. The metal status of their nursing mothers is affected by lifestyle and nutritional factors. However, the relationship between these factors and breastmilk metal concentration is largely unknown.

The present study provided food frequency questionnaires to be completed by 113 nursing mothers from Fujian Provincial Hospital. Their breastmilk samples were analysed for the metal concentrations using inductively coupled plasma mass spectrometery (ICP-MS). Based on the outcome of the questionnaires and ICP-MS results, a total of 94 samples were deemed valid for further data analyses.

Results showed levels of breastmilk lead (Pb) and iron (Fe) were outside the reference range from authoritative data. Correlation analyses found Pb levels to negatively associate with dietary fibre intake. Additionally, Fe levels were negatively associated with alcohol and dairy consumption. In terms of Zn levels, it was positively associated with tea and vitamin B1 intake. Zn was also negatively associated with seafood consumption.

The study concluded dietary factors to associate with metal levels within breastmilk. Recommendations were made to increase consumption of dietary fibre, tea and foods rich in vitamin B1 for gestational and postpartum women. Conversely, alcohol consumption continues to be discouraged for this population. Furthermore, careful considerations need to be taken for levels of dairy and seafood intake to minimize metal metabolism disruptions. Infant metal exposure requires critical attention and this needs to be initiated through the diet and lifestyle of their nursing mothers.

## 1. Introduction

There is a rapid growth and development during infancy until two years of age (1). Metal provides important biological functions during infancy phase (1). It was reported that zinc (Zn) and iron (Fe) play important roles in haematopoiesis, immune function and neurodevelopment (1). Fe deficiency is a prevalent problem worldwide (2). It leads to adverse long term health consequences such as in neuropsychological function (3, 4). Another important metal, calcium (Ca) is the essential building blocks for teeth and bones as well as other roles in muscle contraction, neurotransmission and vascular functions. Conversely, metals may exert detrimental effects on infant development including lead (Pb), cadmium (Cd), Arsenic (As) and aluminium (Al) (5–7).

Since breastmilk is generally the sole food for infants (8), it is a most significant source of these metals (9). Breastfeeding is recommended to be carried out exclusively until six months of infant age and partially until one year of infant age (10). Lactation occurs through endocrine system control of progesterone, oestrogen, prolactin and oxytocin (11). In the early stage of postpartum, breastmilk is produced in a form of colostrum (12), and differentially profiled breastmilk is lactated including foremilk and hindmilk (12). Breastfeeding has been associated with long term obesity prevention, better skeletal outcomes and improved cognition in offsprings (13–15).

Diet contributes as a substantial source of metal intake. Randomized clinical trials have demonstrated beef consumption to improve Fe status in athletes and young women (16, 17). In contrast, harmful metals may also be transferred through dietary sources. This is evident for Cd, Pb and As that have contaminated vegetables or other food agricultural crops in China (18, 19). Few researches have shown the association between nutrient intakes with changes in the breastmilk composition. For example, Ca supplementation in Gambian women was able to improve their breastmilk Ca concentrations (20). Furthermore, multivitamin supplementation improved vitamin E and retinol concentrations in breastmilk of Tanzanian HIV-infected nursing mothers (21).

Other environmental factors are also important influences of metal intake such as the use of public transport and exposure to cigarette smoke (22, 23). Additional studies have demonstrated environmental factors to influence the breastmilk composition. In a study of investigating breastmilk from Brazillian mothers, they found similar metal profiles between their drinking water and their breastmilk (24). In another study, Iranian mothers were analysed for their breastmilk and a variety of environmental factors (25). Their Cd levels appeared higher when they resided near the city waste disposal site. They additionally found unemployed mothers to possess higher Pb levels, possibly due to more frequent Pb exposure from house chores (25).

While breastmilk is imperative for infant health, there is exceedingly limited research surrounding maternal nutrition on breastmilk composition. Deficiency in essential metals and excessive intake of harmful metals during infancy remains a global issue. This needs to be strategized through their nursing mothers. As such, the present study aimed to analyse the relationship between dietary habits and breastmilk metal profile. There were more studies addressing the relationship between lifestyle factors with breastmilk metal profile. Different lifestyle factors were simultaneously assessed using the same cohort of Fuzhou nursing mothers at postpartum period of day 42.

## 2. Methods

### 3.1 Study Subjects

Nursing mothers that seek medical care in Fujian Provincial Hospital (2012 January – April), were selected to investigate their lifestyle, dietary habits and health status. They lived in the Fuzhou area long term. Exclusion criteria included absence of alcohol, tobacco and recent medication consumption. Those with breast, mammary gland, heart, liver, lung, kidney, congenital and genetic diseases were also excluded from the study. The study was approved by the Biomedical research Ethics Committee at Fujian Centre for Disease Control & Prevention.

### 3.2 Food Frequency Questionnaires

Food frequency questionnaires were used for the nursing mothers to record their diet during their first 30 days of their postpartum period (n = 113). The nursing mothers were trained prior to the study including in the project definitions, dietary investigation methods, food and cutlery items, sample collection and clinical data. The questionnaires were modified according to feedbacks given during the training sessions to maximize clarity. The questionnaires were immediately reviewed after retrieval in order for prompt corrections where necessary.

### 3.3 Breastmilk analysis

Their breastmilk samples of 20 mL were collected at postpartum period of day 42. These were stored immediately at −20 °C until analysis (n = 113). 400μL of 69% nitric acid and 400μL of H_2_O_2_ (400μL) were added to 200 μL of breastmilk. Mars-5 microwave dissolver (CEM, Matthews, NC, USA) was used to dissolve the samples and then further diluted down to 5 g using deionized water. The metals were analysed on an Agilent 7500 ce inductively coupled plasma mass spectrometer (Agilent Technologies, Wilmington, DE, USA). These included aluminium (Al), arsenic (As), barium (Ba), calcium (Ca), cadmium (Cd), chromium (Cr), copper (Cu), iron (Fe), potassium (K), magnesium (Mg), manganese (Mn), sodium (Na), nickel (Ni), antimony (Sb), selenium (Se) and zinc (Zn). It was conducted using external standard of certified standard reference material (GBW08509a defatted milk powder, National Research Center for Certified Reference Materials, Beijing, China). Parallel sampling of 10% breastmilk was used to minimize bias from inter-run variation.

### 3.4 Data Analysis

The questionnaires were inspected, coded, assigned and the data was doubly entered (EpiData – *which version*, Odense, Denmark). The entered data were checked repeatedly to ensure the entered data concords with the original data. Based on questionnaire completion and breastmilk abnormalities, 94 samples were used for further analysis.

Statistical analysis was carried out using SPSS 13.0 (IBM, Armonk, NY, US). Normality testing of breastmilk metals found K, Ca, Cu, Mg and Se to be Normal distributed and proceeded with multiple linear regression analysis (α = 0.05). Comparison between single metals were made using analysis of variance (α = 0.05, normal distribution) and non-parametric Mann-Whitney-Wilcoxon test (α = 0.05, non-normal distribution). Pearson’s correlation (normal distributed) and Spearman correlation (non-normal distributed) two-sided tests were also conducted (significance level p < 0.05). The variable assignment of influencing factors presented in Table 1.

**Table 1.** Influencing factors of breastmilk and their variable assignment.

| Influencing factor | 0 | 1 | 2 | 3 |
| --- | --- | --- | --- | --- |
| Geographical location | - | Mountainous | Plains | Coastal |
| Nursing period | - | Beginning | Intermediate | Late |
| Occupation nature | - | Minimal<br>exercise | Moderate<br>exercise | Frequent<br>exercise |
| Monthly income | - | $\leq 4000$ CNY | 4000 - 8000<br>CNY | $> 8000$ CNY |
| Smokers in the family | No | Yes | - | - |
| Alcohol consumption | No | Yes | - | - |
| Tea consumption | No | Yes | - | - |
| Exercise before<br>delivery | No | Yes | - | - |
| Delivery method | - | Vaginal | Assisted vaginal | Caesarean |
| Adequate dairy intake | No | Yes | - | - |
| Adequate meat intake | No | Yes | - | - |

## 3. Results

### 3.1 Demographics

The demographics of the study subjects were summarised in Table 2. On average, the study subjects were 28.5 ± 3.5 years of age and were from the plains-type (Fuzhou five district) geographical location of 74.5%. They were generally educated with an undergraduate degree of 61.7% and with different occupations.

**Table 2.** Demographics of study subjects.

|  |  |
| --- | --- |
| Age | 28.5±3.5 years of age |
| Geographical Location | n = 94 (100%) |
| Mountains (Minho, Yongtai) | 11 (11.7%) |
| Plains (Fuzhou five district) | 70 (74.5%) |
| Coastal (Changle, Fuqing, Lianjiang, Luoyuan, Pingtan) | 13 (13.8%) |
| Education | n = 94 (100%) |
| Secondary or Under | 26 (27.7%) |
| Undergraduate | 58 (61.7%) |
| Postgraduate | 9 (9.6%) |
| Occupation | n = 94 (100%) |
| Company Clerk or Others | 33 (35.1%) |
| Professionals & Civil Service | 30 (31.9%) |
| At-Home | 31 (33.0%) |

### 3.2 Quantity of breastmilk metals

The concentrations of the analysed 17 metals in breastmilk were shown in Table 3. Due to the lack of meta-analyses of breastmilk metals, the present study had chosen more authoritative publications for comparison. These include the 1990 World Health Organization (WHO), International Atomic Energy Agency (IAEA) report of breastmilk metal concentrations and the raw milk contaminant limits from National Food Safety Standards (NFSS) (GB2762 – 2012).

**Table 3.**
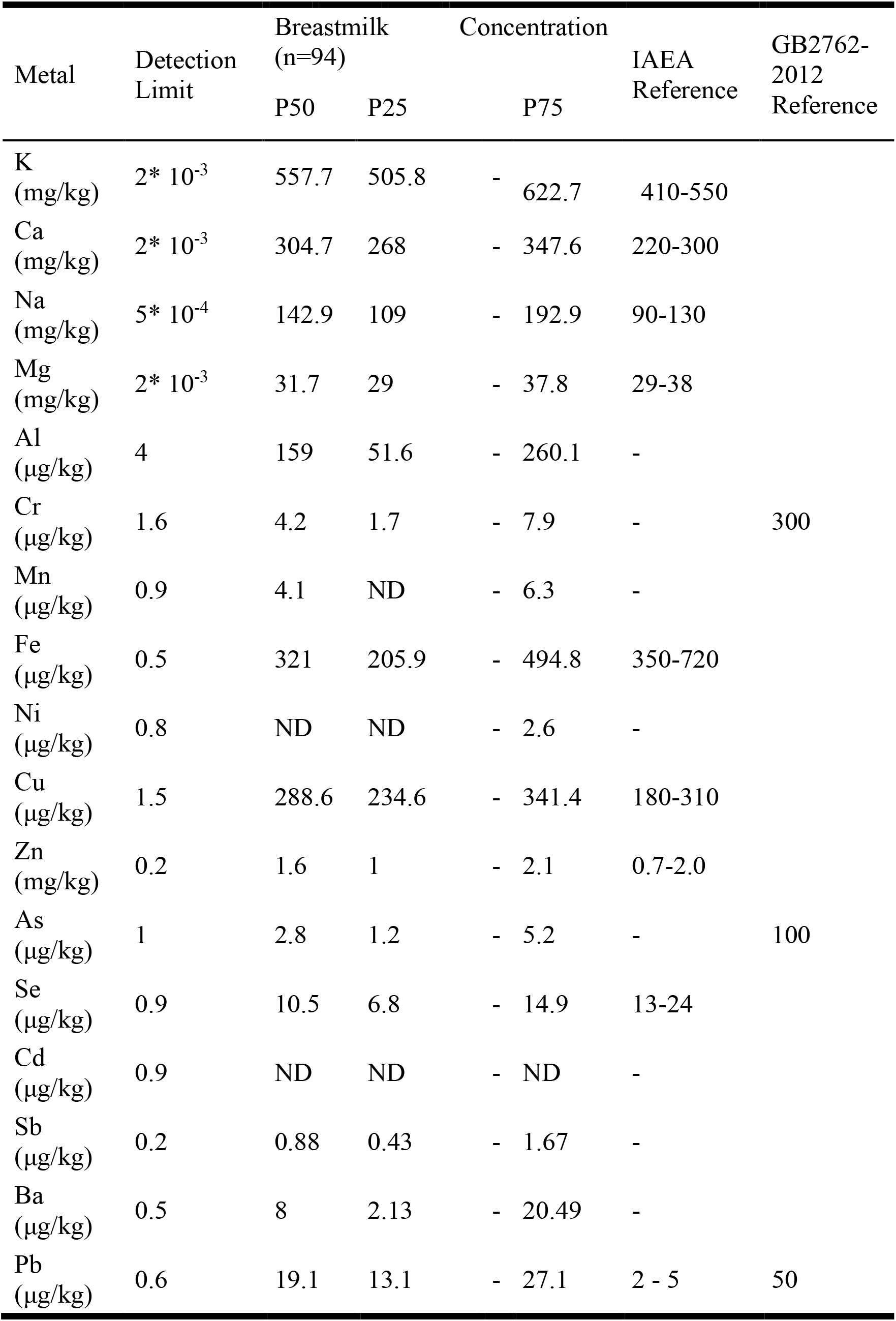
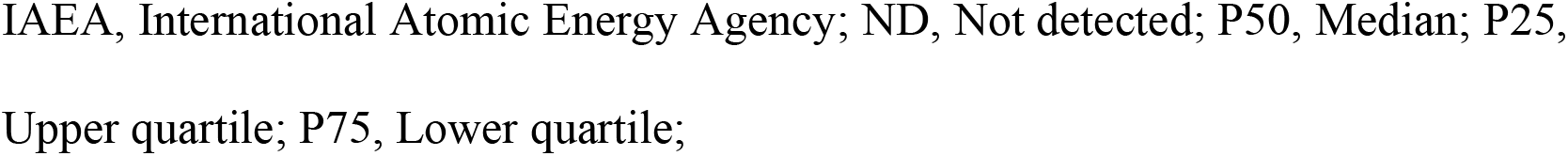
Breastmilk metal concentrations.

| Metal | Detection Limit | Breastmilk (n=94) |  | Concentration |  | IAEA Reference | GB2762-2012 Reference |
| --- | --- | --- | --- | --- | --- | --- | --- |
|  |  | P50 | P25 |  | P75 |  |  |
| K (mg/kg) | $2 \times 10^{-3}$ | 557.7 | 505.8 | - | 622.7 | 410-550 | |
| Ca (mg/kg) | $2 \times 10^{-3}$ | 304.7 | 268 | - | 347.6 | 220-300 | |
| Na (mg/kg) | $5 \times 10^{-4}$ | 142.9 | 109 | - | 192.9 | 90-130 | |
| Mg (mg/kg) | $2 \times 10^{-3}$ | 31.7 | 29 | - | 37.8 | 29-38 | |
| Al (µg/kg) | 4 | 159 | 51.6 | - | 260.1 | - |  |
| Cr (µg/kg) | 1.6 | 4.2 | 1.7 | - | 7.9 | - | 300 |
| Mn (µg/kg) | 0.9 | 4.1 | ND | - | 6.3 | - |  |
| Fe (µg/kg) | 0.5 | 321 | 205.9 | - | 494.8 | 350-720 |  |
| Ni (µg/kg) | 0.8 | ND | ND | - | 2.6 | - |  |
| Cu (µg/kg) | 1.5 | 288.6 | 234.6 | - | 341.4 | 180-310 |  |
| Zn (mg/kg) | 0.2 | 1.6 | 1 | - | 2.1 | 0.7-2.0 |  |
| As (µg/kg) | 1 | 2.8 | 1.2 | - | 5.2 | - | 100 |
| Se (µg/kg) | 0.9 | 10.5 | 6.8 | - | 14.9 | 13-24 |  |
| Cd (µg/kg) | 0.9 | ND | ND | - | ND | - |  |
| Sb (µg/kg) | 0.2 | 0.88 | 0.43 | - | 1.67 | - |  |
| Ba (µg/kg) | 0.5 | 8 | 2.13 | - | 20.49 | - |  |
| Pb (µg/kg) | 0.6 | 19.1 | 13.1 | - | 27.1 | 2 - 5 | 50 |

IAEA, International Atomic Energy Agency; ND, Not detected; P50, Median; P25, Upper quartile; P75, Lower quartile;

In comparison to the IAEA report, Mg, Cu and Zn were within normal concentrations. K, Na and Ca were slightly higher than normal, but Pb was four times higher. Fe and Se were found to be lower. In reference to the NFSS, Cr, As and Pb were lower than the limit.

### 3.3 Influencing factors of normal distributed breastmilk metals

Ca concentration was influenced by the delivery method, with vaginal > assisted vaginal > Caesarean (b’ = −0.267, p<0.05). Subjects with sufficient dairy intake or meat intake (according to nursing mothers food pyramid) (26) had higher Cu (b’ = 0.300; p<0.01) and Se (b’=0.213, p<0.05) levels, respectively. None of the selected independent variables had collinearity between them (variance inflation factor <5).

### 3.4 Influencing factors of non-normal distributed breastmilk metals

The level of element As in the breastmilk differed with geographical location (p<0.05). The exposure to smoking environment for more than 15 minutes per day associated with lower K levels and higher Al and Cd levels (p<0.05). Alcohol and tea consumption prior to gestation associated with lower Fe (p<0.05) and higher Zn (p<0.05) levels, respectively. Adequate intake of fruits, dairy and vitamin B_1_ were found to associate with higher Sb (p<0.05), lower Ni (p<0.05) and higher Zn (p<0.05) levels, respectively.

### 3.5. Correlation analysis of breastmilk metals and diet

The present study found negative correlation between seafood intake with Ca (r = −0.146, p<0.05) and Zn levels (r = −0.166, p<0.05). There was a positive correlation with egg consumption and Mn (r = 0.170, p<0.05). Cu was correlated with seafood intake (r = −0.163, p<0.05), dairy consumption (r = 0.156, p<0.05) and Fe consumption (r = −0.141, p<0.05). Sb was correlated with fruit consumption (r = 0.165, p<0.05), vitamin E intake (r = 0.148, p<0.05) and Vitamin B_12_ intake (r = 0.147, p<0.05).

## 4. Discussion

The importance of metals for infant development is unambiguous. These include Fe for neurodevelopment (27), Zn for immunity (28), Ca for bone mineralization (29) and Cu for cardiovascular health (30). Conversely, contaminant metals can lead to adverse events in the infant such as alterations in memory by excessive Pb (31). Due to their sole food being generally breastmilk during infnancy (8), it is therefore the predominant source of these metals (9). Retrospectively, the exposure of these metals in the nursing mothers primarily derives from the external environment through means of diet (18, 19) and lifestyle (22, 23).

There were few studies conducted to evaluate how whole diet influence the metal concentrations within breastmilk. Existing studies has been focused on nutraceutical supplementation (20, 32). There were more studies that investigated how lifestyle affects breastmilk metal concentrations (24, 25). In the present study, we investigated how dietary habits related to the metal profile concentrations in the breastmilk of nursing mothers. In complementary to this, their lifestyles were also evaluated. Since breastmilk exposes infants to a wide range of both physiologically beneficial and adverse metals, this study allows better optimization of infant health. This could be facilitated through improved lifestyle and dietary recommendations to both gestational and postpartum mothers.

In comparison to the IAEA report, the Pb levels of 19.1 μg/kg found in the breastmilk of the present study exceeded the reference range by approximately four times (2 - 5 μg/kg). In aging primates, the infant exposure to Pb reprogrammed a wide array of their gene expression through epigenetics (33). These genes contributed to increased propensity of neurodegenerative diseases such as Alzheimer’s disease. In terms of a more immediate impact, the *in utero* exposure to Pb was associated with lower overall, cognitive and language scores of the “Comprehensive Developmental Inventory for Infants and Toddlers” at two years of age (34). Other breastmilk studies also revealed excessive Pb levels of a shocking 40.6 μg/kg from China (35) and 40.6 μg/kg from Mexico (36). These demonstrated that excess amount of Pb in breastmilk may be a prevalent issue in China and also of substantial global concern.

The present study found negative associations of Pb with dietary fibre intake. This relationship may relate to the influence of dietary fibre on Pb absorption, retention and excretion. It has been demonstrated that dietary fibre can modify tissue retention of Pb in male Lister rats with varying degrees with different types of dietary fibre (37). Recently, pectin and alginate-based dietary fibres were found to bind to Pb, preventing their gastrointestinal absorption and tissue accumulation, as well as increasing their faecal excretion in male Wistar rats (38).

The Fe levels of 321 μg/kg in the present study were below the reference range recommended by the IAEA report of 350 – 750 μg/kg. Fe deficiency during infancy is known to lead to adverse health outcomes in the long term (3, 4). In one study involving Chilean children, Fe deficiency anaemia at six or twelve months into infancy was associated with neuroendocrine changes in cortisol response at ten years of age (4). A more recent study by the same research group found Fe deficiency during infancy was associated with poorer psychological health and educational achievements at 25 years of age (3). The prevalence of iron deficiency in breastfed infants differs in different parts of the world, such as 6% in Sweden, 17% in Mexico and as high as 37% in Ghana (2). These figures in combination with our results express the need to address this Fe deficiency problem at the global level.

Alcohol consumption was found to negatively associate with Fe levels. This result is in disagreement with all of the previous studies which found alcohol consumption to increase the Fe levels (39–42). The molecular mechanisms underlying this effect remain to be fully elucidated. Emerging evidence suggested that alcohol consumption up-regulates iron transporter in the intestinal epithelium (42). This discrepancy with our results may relate to differences in the study subjects. We studied exclusively on postpartum nursing mothers, which lie in the perinatal period that is known to possess unique Fe metabolism (43). Nevertheless, it is strongly prohibited to consume alcohol to improve Fe status as alcohol causes many other adversities to the infant that outlies the benefits of sufficient Fe status (44).

In addition to this association, dairy consumption was also negatively associated with Fe levels. No studies exist to date that have reported on similar findings. However, there is substantial evidence that consumption of cow’s milk was strongly associated with diminished Fe stores in infants (45). This association was speculated to be due to the high proportion of Ca and casein, both of which have shown to interfere with Fe absorption (45). Ca is a known inhibitor of Fe absorption (46). Cow’s milk with higher Ca content has clinically shown to attribute to lower bioavailability of Fe (46). In another clinical trial, casein was found to lower Fe bioavailability when it substituted with egg white protein (47). It is therefore important to balance dairy consumption during postpartum period to minimize Fe interference but simultaneously obtain sufficient benefits from dairy products. Consumption of dairy products remains important as they provide a range of health benefits (48).

Zn levels in our breastmilk samples were observed to associate with higher tea consumption. This observation concords with a study where green tea decoction increased Zn levels in the blood and multiple tissues by using a rat model (49). Green tea extract supplementation has shown similar association in serum samples of obese patients (50). These effects have been hypothesized to attribute to the polyphenol interactions with Zn that influences their absorption (51) or the improved antioxidant status in the body (50). In old Sprague-Dawley rats, effects of tea consumption on Zn absorption depend on the type of tea (51). However, other related studies failed to find the presence of these effects (52–55) and therefore warrant further investigation.

Higher vitamin B1 intake was associated with higher Zn levels. This possibly arise from the simultaneous consumption of foods rich in both Zn and vitamin B1. For example, 100 g of pork may contribute 33% daily intake of Zn and 50% daily intake of vitamin B1 (56–58). Mung beans are also excellent sources of Zn and vitamin B1 with 100 g contributing to 6% daily intake of Zn and 10% daily intake of vitamin B1 (59, 60). Both of these nutrients are critically important during infancy (61–63). Zn is involved in protein synthesis and other cellular processes (61). Vitamin B1 deficiency causes beriberi, which is characterized with cardiovascular adversities (62).

The present study discovered seafood to negatively associate with Zn levels. This was a surprising finding because it is known that seafoods such as oyster, scallop, crabs, acts as a rich source of Zn (64, 65). However, no existing related studies have been found. Furthermore, other types of seafoods were found to be less rich in Zn including pout, paella and gurnard (64). Therefore, further investigation into seafood subtypes may be useful. The apparent negative association may relate to seafood nutrient interactions with Zn metabolism. For example, it has been demonstrated that many different types of protein affects Zn bioavailability (66–68). Given the negative association, it is important to balance sufficient Zn intake without abstaining from seafood because seafood is known to provide a wide range of benefits perinatally (69).

There were several limitations within the present study. The breastmilk samples were taken at a single postpartum time point, which may not be representative of its metal concentrations in the extended postpartum period. Furthermore, only Fuzhou subjects that visited the Fujian Provincial Hospital were recruited. This reduces the translation of our findings to the entire Fuzhou, greater China and at the global level. The sample size of the study was too small to conclude on any associations observed. In saying that, this research still presents value due to the lack of breastmilk nutrition research and the critical global concern for metal intake during infancy. The observed associations provided entry for further conclusive research to be made. The restricted origin of the study subjects provided a specifically defined cohort that allowed meaningful breastmilk data to be compiled. Having samples from mixed regions could create substantial bias due to the significant influence of environment on breastmilk characteristics (70). The results draws attention to possible existing breastmilk issues within the region that can inspire further research to be translated to a wider population.

In future studies, multiple breastmilk samples across the postpartum period would be beneficial to evaluate the results or trends. This can be complemented by the physiological parameters of the infant for correlations to be made or metabolic adaptations to be identified. In order to attain results with higher robustness, the sample size needs to be increased. The Fuzhou population can be more represented if future studies involve multiple Fuzhou hospitals in both rural and urban areas. The associations found in the present study should be explored further in more details. This includes higher characterization of dietary components or carrying out randomized clinical trials. With a more targeted focus, multiple types of measurements of metals can reduce impact from weakness of particular experimental methodologies.

## 5. Conclusion

The present study addressed the breastmilk metal status in Fuzhou nursing mothers at postpartum period of 42 days. Multiple associations were made for these with lifestyle and dietary factors. It was discovered that excess Pb associated negatively with dietary fibre intake. Therefore, it is recommended to increase dietary fibre intake that may decrease adverse Pb transfer from breastmilk to the infants. Additionally, there appears to be a state of Fe deficiency and these levels were negatively associated with alcohol and dairy consumption. Thus, care should be taken to avoid Fe deficiency. This could be improved through abstinence from alcoholic beverages and regarding the risk-benefit balance for dairy products.

In terms of the Zn levels, positive associations were found with tea and vitamin B1 intake as well as negative associations with seafood. These findings encourage drinking tea and consuming foods that are rich in Zn and vitamin B1 (e.g. pork, mung beans). Conversely, seafood should be consumed in moderation by judging between its derived benefits and possible adverse effects on Zn metabolism.

The present study addressed research gaps surrounding breastmilk metal levels with maternal dietary factors. This is an important topic at the global level because of breastmilk being the predominant source of nutrients for infants. Heavy metal intake and deficiency of essential metals during infancy are a worldwide concern due to its prevalence and its long term impact into adulthood. By understanding the nutritional contribution to breastmilk metals in nursing mothers, adverse events can be minimized for the infants during breastfeeding in order to optimize their developmental potential.

## Data Availability

The datasets generated during and/or analysed during the current study are available from the corresponding author on reasonable request.

## Abbreviations

IAEA: International Atomic Energy Agency
NFSS: National Food Safety Standards
Al: Aluminium
As: Arsenic
Ba: Barium
Ca: Calcium
Cd: Cadmium
Cr: Chromium
Cu: Copper
Fe: Iron
K: Potassium
Mg: Magnesium
Mn: Manganese
Na: Sodium
Ni: Nickel
Sb: Antimony
Se: Selenium
Zn: Zinc

## Acknowledgements

The authors would like to thank the chief physician, Pin Ge and physician, Guo-Bo Li from Fujian Province Maternity and Child Care Center, Fujian Province Chinldren’s hospital for their assistance to the sample and questionnaire collection. The authors would like to thank the senior technologist, Guo-Bin Lin from Fujian Center for Disease Control &Prevention for his assistance with sample testing and data analysis. The authors would like to thank the participation of the postpartum women.

## References

1. Hermoso M, Tabacchi G, Iglesia-Altaba I, Bel-Serrat S, Moreno-Aznar LA, Garcia-Santos Y, et al. The nutritional requirements of infants.towards EU alignment of reference values: The EURRECA network. Matern Child Nutr. 2010;6:55–83.

2. Yang Z, Lonnerdal B, Adu-Afarwuah S, Brown KH, Chaparro CM, Cohen RJ, et al. Prevalence and predictors of iron deficiency in fully breastfed infants at 6 months of age: Comparison of data from 6 studies. Am J Clin Nutr. 2009;89:1433–1440.

3. Lozoff B, Smith JB, Kaciroti N, Clark KM, Guevara S, Jimenez E. Functional significance of early-life iron deficiency: Outcomes at 25 years. J Pediatr. 2013;163(5):1260–1266.

4. Felt BT, Peirano P, Algarin C, Chamorro R, Sir T, Kaciroti N, et al. Long-term neuroendocrine effects of iron-deficiency anemia in infancy. Pediatr Re. 2012;71(6):707–712.

5. Kim Y, Ha EH, Park H, Ha M, Kim Y, Hong YC, et al. Prenatal lead and cadmium co-exposure and infant neurodevelopment at 6 months of age: The mothers and children’s environmental health (MOCEH) study. NeuroToxicology. 2013;35:15–22.

6. Sakamoto M, Chan HM, Domingo JL, Kubota M, Murata K. Changes in body burden of mercury, lead, arsenic, cadmium and selenium in infants during early lactation in comparison with placental transfer. Ecotoxicol Environ Saf. 2012;84:179–184.

7. Dabeka R, Fouquet A, Belisle S, Turcotte S. Lead, cadmium and aluminum in canadian infant formulae, oral electrolytes and glucose solutions. Food Addit Contam. 2011;28(6):744–753.

8. Johns HM, Forster DA, Amir LH, McLachlan HL. Prevalence and outcomes of breast milk expressing in women with healthy term infants: A systematic review. BMC Pregnancy Childbirth. 2013;13:212.

9. Gidrewicz DA, Fenton TR. A systematic review and meta-analysis of the nutrient content of preterm and term breast milk. BMC Pediatr. 2014;14:216.

10. Hunter T, Cattelona G. Breastfeeding initiation and duration in first-time mothers: Exploring the impact of father involvement in the early post-partum period. Health Promot Perspect. 2014;4(2):132–136.

11. Stuebe AM, Grewen K, Pedersen CA, Propper C, Meltzer-Brody S. Failed lactation and perinatal depression: Common problems with shared neuroendocrine mechanisms? J Womens Health. 2012;21(3):264–272.

12. Mangel L, Ovental A, Batscha N, Arnon M, Yarkoni I, Dollberg S. High fat content in breastmilk expressed manually: A randomized trial. Breastfeed Med.2015.

13. Moss BG, Yeaton WH. Early childhood healthy and obese weight status: Potentially protective benefits of breastfeeding and delaying solid foods. Matern Child Health J. 2014;18:1224–1232.

14. Jones G, Hynes KL, Dwyer T. The association between breastfeeding, maternal smoking in utero, and birth weight with bone mass and fractures in adolescents: A 16-year longitudinal study. Osteoporos Int. 2013;24:1605–1611.

15. Belfort MB, Rifas-Shiman SL, Kleinman KP, Guthrie LB, Bellinger DC, Taveras EM, et al. Infant feeding and childhood cognition at ages 3 and 7 years: Effects of breastfeeding duration and exclusivity. JAMA Pediatr. 2013;167(9):836–844.

16. Blanton C. Improvements in iron status and cognitive function in young women consuming beef or non-beef lunches. Nutrients. 2014;6:90–110.

17. Burke DE, Johnson JV, Vukovich MD, Kattelmann KK. Effects of lean beef supplementation on iron status, body composition and performance of collegiate distance runners. Food Nutr Sci. 2012;3:810–821.

18. Huang Z, Pan XD, Wu PG, Han JL, Chen Q. Heavy metals in vegetables and the health risk to population in zhejiang, china. Food Control. 2014;36:248–252.

19. Zhao Y, Fang X, Mu Y, Cheng Y, Ma Q, Nian H, et al. Metal pollution (cd, pb, zn, and as) in agricultural soils and soybean, glycine max, in southern china. Bull Environ Contam Toxicol. 2014;92:427–432.

20. Jarjou LMA, Prentice A, Sawo Y, Laskey MA, Bennett J, Goldberg GR, et al. Randomized, placebo-controlled, calcium supplementation study in pregnant gambian women: Effects on breast-milk calcium concentrations and infant birth weight, growth, and bone mineral accretion in the first year of life. Am J Clin Nutr. 2006;83:657–666.

21. Webb AL, Aboud S, Furtado J, Murrin C, Campos H, Fawzi WW, et al. Effect of vitamin supplementation on breast milk concentrations of retinol, carotenoids and tocopherols in HIV-infected tanzanian women. Eur J Clin Nutr. 2009;63:332–339.

22. Gao P, Liu S, Ye W, Lin N, Meng P, Feng Y, et al. Assessment on the occupational exposure of urban public bus drivers to bioaccessible trace metals through resuspended fraction of settled bus dust. Sci Total Environ. 2015;508:37–45.

23. Rzymski P, Rzymski P, Tomczyk K, Niedzielski P, Jakubowski K, Poniedzialek B, et al. Metal status in human endometrium: Relation to cigarette smoking and histological lesions. Environ Res. 2014;132:328–333.

24. Cardoso OO, Juliao FC, Alves RIS, Baena AR, Diez IG, Suzuki MN, et al. Concentration profiles of metals in breast milk, drinking water, and soil: Relationship between matrices. Biol Trace Elem Res. 2014;160:116–122.

25. Dursun A, Yurdakok K, Yalcin SS, Tekinalp G, Aykut O, Orhan G, et al. Maternal risk factors associated with lead, mercury and cadmium levels in umbilical cord blood, breast milk and newborn hair. J Matern Fetal Neonatal Med. 2015:1476–7058.

26. Chinese Nutrition Society. Dietary guidelines for chinese gestational and breastfeeding women and children 0 - 6 years of age. 2007th ed. Beijing, China: People’s Medical Publishing House; 2007.

27. Domellof M, Braegger C, Campoy C, Colomb V, Decsi T, Fewtrell M, et al. Iron requirements of infants and toddlers. J Pediatr Gastroenterol Nutr. 2014;58(1):119–129.

28. Abrams SA. Zinc for preterm infants: Who needs it and how much is needed? Am J Clin Nutr. 2013;98:1373–1374.

29. Trindade CEP. Minerals in the nutrition of extremely low birth weight infants. J Pediatr. 2005;81(1):S43–S51.

30. Bhatia J, Griffin I, Anderson D, Kler N, Domellof M. Selected macro/micronutrient needs of the routine preterm infant. J Pediatr. 2013;162(3):S48–S55.

31. Geng F, Mai X, Zhan J, Xu L, Shao J, Meeker J, et al. Low-level prenatal lead exposure alters auditory recognition memory in 2 month old infants: An event-related potentials (ERPs) study. Dev Neuropsychol. 2014;39(7):516–528.

32. Sazawal S, Black RE, Dhingra P, Jalla S, Krebs N, Malik P, et al. Zinc supplementation does not affect the breast milk zinc concentration of lactating women belonging to low socioeconomic population. J Hum Nutr Food Sci. 2013;1(2):1014.

33. Bihagi SW, Huang H, Wu J, Zawia NH. Infant exposure to lead (pb) and epigenetic modifications in the aging primate brain: Implications for alzheimer’s disease. J Alzheimers Dis. 2011;27(4):819–833.

34. Lin CC, Chen YC, Su FC, Lin CM, Liao HF, Hwang YH, et al. In utero exposure to environmental lead and manganese and neurodevelopment at 2 years of age. Environ Res. 2013;123:52–57.

35. Liu K, Hao J, Xu Y, Gu X, Shi J, Dai C, et al. Breast milk lead and cadmium levels in suburban areas of nanjing, china. Chin Med J. 2013;28(1):7–15.

36. Amarasiriwardena CJ, Jayawardene I, Lupoli N, Barnes RM, Hernandez-Avila M, Hu H, et al. Comparison of digestion procedures and methods for quantification of trace lead in breast milk by isotope dilution inductively coupled plasma mass spectrometry. Anal Methods. 2013;5:1676.

37. Rose HE, Quarterman J. Dietary fibers and heavy metal retention in the rat. Environ Res. 1987;42:166–175.

38. Khotimchenko M, Serguschenko I, Khotimchenko Y. Lead absorption and excretion in rats given insoluble salts of pectin and alginate. Int J Toxicol. 2006;25:195–203.

39. Ioannou GN, Dominitz JA, Weiss NS, Hegarty PJ, Kowdley KV. The effect of alcohol consumption on the prevalence of iron overload, iron deficiency, and iron deficiency anemia. Gastroenterology. 2004;126:1293–1301.

40. Whitfield JB, Zhu G, Heath AC, Powell LW, Martin NG. Effects of alcohol consumption on indices of iron stores and of iron stores on alcohol intake markers. Alcohol Clin Exp Res. 2001;25(7):1037–1045.

41. Chapman RW, Morgan MY, Laulicht M, Hoffbrand AV, Sherlock S. Hepatic iron stores and markers of iron overload in alcoholics and patients with idiopathic hemochromatosis. Dig Dis Sci. 1982;27(10):909–916.

42. Harrison-Findik DD. Role of alcohol in the regulation of iron metabolism. World J Gastroenterol. 2007;13(37):4925–4930.

43. Koenig MD, Tussing-Humphreys L, Day J, Cadwell B, Nemeth E. Hepcidin and iron homeostasis during pregnancy. Nutrients. 2014;6:3062–3083.

44. O’Leary CM, Jacoby PJ, Bartu A, D’Antoine H, Bower C. Maternal alcohol use and sudden infant death syndrome (SIDS) and infant mortality excluding SIDS. Pediatrics. 2013;131(3):e770–e778.

45. Ziegler EE. Consumption of cow’s milk as a cause of iron deficiency in infants and toddlersnure. Nutr Rev. 2011;69(1):S37–S42.

46. Hallberg L, Rossander-Hulten L, Brune M, Gleerup A. Bioavailability in man of iron in human milk and cow’s milk in relation to their calcium contents. Pediatr Res. 1992;31(5):524–527.

47. Hurrell RF, Lynch SR, Trindad TP, Dassenko SA, Cook JD. Iron absorption in humans as influenced by bovine milk proteins. Am J Clin Nutr. 1989;49:546–552.

48. Brantsaeter AL, Olafsdottir AS, Forsum E, Olsen SF, Thorsdottir I. Does milk and dairy consumption during pregnancy influence fetal growth and infant birthweight? A systematic literature review. Food Nutr Res. 2012;56.

49. Hamdaoui MH, Chahed A, Ellouze-Chabchoub S, Marouani N, Ben Abid Z, Hedhili A. Effect of green tea decoction on long-term iron, zinc and selenium status of rats. Ann Nutr Metab. 2005;49:118–124.

50. Suliburska J, Bogdanski P, Szulinska M, Stepien M, Pupek-Musialik D, Jablecka A. Effects of greeen tea supplementation on elements, total antioxidants, lipids, and glucose values in the serum of obese patients. Biol Trace Elem Res. 2012;149:315–322.

51. Deng Z, Tao B, Li X, He J, Chen Y. Effect of green tea and black tea on the metabolisms of mineral elements in old rats. Biol Trace Elem Res. 1998;65:75–86.

52. Basu A, Betts NM, Mulugeta A, Tong C, Newman E, Lyons TJ. Green tea supplementation increases glutathione and plasma antioxidant capacity in adults with the metabolic syndrome. Nutr Res. 2013;33(3):180–187.

53. Ganji V, Kies CV. Zinc bioavailability and tea consumption. Plant Foods Hum Nutr. 1994;46:267–276.

54. Perez-Llamas F, Gonzalez D, Cabrera L, Espinosa C, Lopez JA, Larque E, et al. White tea consumption slightly reduces iron absorption but not growth, food efficiency, protein utilization, or calcium, phosphorus, magnesium, and zinc absorption in rats. J Physiol Biochem. 2011;67:331–337.

55. Record IR, McInernery JK, Dreosti IE. Black tea, green tea, and tea polyphenols. effects on trace element status in weanling rats. Biol Trace Elem Res. 1996;53:27–43.

56. Lombardi-Boccia G, Lanzi S, Aguzzi A. Aspects of meat quality: Trace elements and B vitamins in raw and cooked meats. J Food Compost Anal. 2005;18(1):39–46.

57. Lombardi-Boccia G, Lanzi S, Lucarini M, di Lullo G. Meat and meat products consumption in italy: Contribution to trace elements, heme iron and selected B vitamins supply. Int J Vitam Nutr Res. 2004;74(4):247–251.

58. Murphy MM, Spungen JH, Bi X, Barraj LM. Fresh and fresh lean pork are substantial sources of key nutrients when these products are consumed by adults in the united states. Nutr Res. 2011;31(10):776–783.

59. Ejigui J, Savoie L, Marin J, Desrosiers T. Influence of traditional processing methods on the nutritional composition and antinutritional factors of red peanuts (arachis hypogea)and small red kidney beans (phaseolus vulgaris). J Biol Sci. 2005;5(5):597–605.

60. Alonso R, Rubio LA, Muzqui M, Marzo F. The effect of extrusion cooking on mineral bioavailability in pea and kidney bean seed meals. Anim Feed Sci Technol. 2001;94:1–13.

61. Chaffee BW, King JC. Effect of zinc supplementation on pregnancy and infant outcomes: A systematic review. Paediatr Perinat Epidemiol. 2012;26(1):118–137.

62. Porter SG, Coats D, Fischer PR, Ou K, Frank EL, Sreang P, et al. Thiamine deficiency and cardiac dysfunction in cambodian infants. J Pediatr. 2014;164(6):1456–1461.

63. Bowman BA, Pfeiffer CM, Barfield WD. Thiamine deficiency, beriberi, and maternal and child health: Why pharmacokinetics matter. Am J Clin Nutr. 2013;98:635–6336.

64. Guerin T, Chekri R, Vastel C, Sirot V, Volatier JL, Leblanc JC, et al. Determination of 20 trace elements in fish and other seafood from the french market. Food Chem. 2011;127:934–942.

65. Bilandzic N, Sedak M, Dokic M, Varenina I, Kolanovic BS, Bozic D, et al. Determination of zinc concentrations in foods of animal origin, fish and shellfish from croatia and assessment of their contribution to dietary intake. J Food Compost Anal. 2014;35(2):61–66.

66. Hemalatha S, Gautam S, Platel K, Srinivasan K. Influence of exogenous iron, calcium, protein and common salt on the bioaccessibility of zinc from cereals and legumes. J Trace Elem Med Biol. 2009;23:75–83.

67. Etcheverry P, Hawthorne KM, Liang L, Abrams SA, Griffin IJ. Effect of beef and soy proteins on the absorption of non-heme iron and inorganic zinc in children. J Am Coll Nutr. 2006;25(1):34–40.

68. Rosado JL, Diaz M, Gonzalez K, Griffin I, Abrams SA, Preciado R. The addition of milk or yoghurt to a plant-based diet increases zinc bioavailability but does not affect iron bioavailability in women. J Nutr. 2005;135:465–468.

69. Brantsaeter AL, Birgisdottir BE, Meltzer HM, Kvalem HE, Alexander J, Magnus P, et al. Maternal seafood consumption and infant birth weight, length and head circumference in the norwegian mother and child cohort study. Br J Nutr. 2012;107:436–444.

70. Lipkie TE, Morrow AL, Jouni ZE, McMahon RJ, Ferruzzi MG. Longitudinal survey of carotenoids in human milk from urban cohorts in china, mexico, and the USA. PLoS ONE. 2015;10(6):e0127729.

